# Judged by your neighbors: A novel framework for personalized assessment of brain structural aging effects in diverse populations

**DOI:** 10.1101/2024.12.24.24319598

**Authors:** Ramona Leenings, Nils R. Winter, Jan Ernsting, Maximilian Konowski, Vincent Holstein, Susanne Meinert, Jennifer Spanagel, Carlotta Barkhau, Lukas Fisch, Janik Goltermann, Malte F. Gerdes, Dominik Grotegerd, Elisabeth J. Leehr, Annette Peters, Lilian Krist, Stefan N. Willich, Tobias Pischon, Henry Völzke, Johannes Haubold, Hans-Ulrich Kauczor, Thoralf Niendorf, Maike Richter, Udo Dannlowski, Klaus Berger, Xiaoyi Jiang, James Cole, Nils Opel, Tim Hahn, NAKO consortium, ADNI consortium, Frontotemporal Lobar Degeneration Neuroimaging Initiative, Australian Imaging Biomarkers, Biomarkers and Lifestyle flagship study of ageing

## Abstract

**Background:** Despite established consensus about the individuality of brain physiology and structural changes, current neuroimaging biomarker paradigms heavily rely on population averages. Here, we define normativity not as a single reference point but as a landscape of permissible configurations arising from diverse physiological manifestations.

**Methods:** We introduce the Nearest Neighbor Normativity (N^3^) framework, designed to parse the natural heterogeneity in brain structural measurements into a diversity-aware biomarker of brain structural decline. Through repeated comparisons within different demographic subgroups, N^3^ contextualizes an individual’s measurements from several viewpoints along the aging continuum. Relying on local density estimation, it accommodates multiple, shared normative configurations within and across age groups. We use T1-weighted MRI data from 29,883 individuals for training and 7,013 individuals for validation, and benchmark our framework against Brain Age models and Normative Modeling. We evaluate each biomarker’s ability to detect neurodegenerative diseases, using them as representative models of atypical brain structural decline.

**Results:** In group-level analyses, the N^3^ biomarker shows the largest effect sizes in the detection of Mild Cognitive Impairment, Alzheimer’s Disease, and Frontotemporal Dementia. Similarly, the N^3^ biomarker achieves the highest predictive performance in the classification of neurodegenerative diseases on a single subject level. Notably, the N^**3**^ estimates remain stable across age and robust to compositional changes in the reference sample. Finally, correlation analyses indicate that it captures distinct and complementary aspects of brain structural variation compared to existing approaches.

**Conclusions:** Our findings underscore that rethinking normativity assessments to explicitly accommodate natural inter-individual variability can enhance the efficacy and translational value of neuroimaging biomarkers and advance our collective efforts toward truly personalized patient care.

## 1 Introduction

Finding (subtle) individual norm deviations moderates our ability to diagnose and treat diseases effectively and is a practical necessity for the personalization of patient care. To this end, neuroimaging biomarkers hold the promise of providing objective indices of neurophysiological health [1–6]. They are closely tied to the concept of brain structural normativity, i.e. the degree of alignment with norms seen in the general population. Despite established consensus about the individuality in brain structure and aging patterns, however, current neuroimaging biomarkers heavily rely on population averages in their quantifications. They thereby exclude the possibility of multiple, equally viable normative health states, potentially obscuring tailored medical interventions to individual physiology [7, 8]. Redefining neuroimaging biomarkers is thus not merely an academic exercise, but holds profound implications for medical practice.

Two prominent approaches have emerged in the search for reliable neuroimaging biomarkers in brain structure thus far. The first approach, called normative modeling, uses statistical distributions to quantify brain structural measures in relation to the population average and the variance around it [9, 10]. It has been successfully applied to detect brain structural norm deviations in various psychiatric disorders [11]. By design, normative models interpolate natural variability into a single reference distribution, centered around the population average. This mean-centric framework neglects the heterogeneity of physiological manifestations and does not account for biologically valid alternative norms. Thus, these population-wide, and often univariate, models risk masking subtle details that are critical for obtaining nuanced individual clinical insights [12].

The second approach, called Brain Age [13, 14], is a derivative of normative modeling and trains machine learning models to predict chronological age from brain structure using examples of a healthy reference sample. The resulting biomarker, the Brain Age Gap (BAG), is defined as the difference between chronological and predicted age and has been statistically associated with numerous neurological and psychiatric conditions, such as Alzheimer’s Disease (+5-10 years), Mild Cognitive Impairment, (+1-10 years), Major Depressive Disorder (BAG 1-4 years), Schizophrenia (+3-12 years). [15–19].

However, the BAG’s ability to capture individual norm deviations is naturally limited due to the multifaceted nature of individual aging processes. How quickly aging effects progress in individuals is highly variable and a complex interplay of genetic predisposition, behavioral choices and cumulative impact of various adverse or protective exposures [20–24]. This unique combination of factors inevitably results in different aging effects seen among same-aged individuals. Using chronological age to assess normative aging effects thus inherently lacks precision. Consequently, the resolution of the BAG as a personalized brain structural assessment tool is limited and its utility mainly constrained to group comparisons.

To address the methodological shortcomings of the approaches described above, we propose a novel normativity estimation framework called Nearest Neighbor Normativity (N^3^).

Normativity in our framework is defined as a structured space of permissible configurations that arises from the diverse physiological manifestations observed in healthy individuals. Healthy inter-individual variability reflects biologically principled movement in this space, whereas anomalies correspond to configurations that deviate in biologically unexpected directions. A biomarker should be able to distinguish physiologically coherent variants from truly atypical patterns that depart from the manifold of healthy configurations. Grounded within this conceptualization, the N^3^ framework shifts from estimating distance to an average, prototypical observation toward locating each individual within a high-dimensional landscape of plausible and implausible configurations.

In this work, we introduce the N^3^ framework and use it to assess the normativity of brain structures while explicitly accounting for the natural and systematic changes along the aging continuum. By relying on context-enriched assessments, the N^3^ is inherently designed to improve the understanding of variance in-between individual brain structures within the natural diversity seen along the aging continuum. We benchmark the resulting N^3^ biomarker relative to conventional neuroimaging biomarkers, including conventional normative modeling and Brain Age models. As a representative use case, we focus on neurodegenerative disorders, which are characterized by structural alterations and deviations from normative aging trajectories, and therefore provide a suitable testbed for the evaluation of the proposed framework.

## 2 Methods

### 2.1 N^3^ algorithm

The N^3^ framework evaluates medical measurements within intentionally selected subgroups of a reference dataset to realize comparisons from multiple, clinically meaningful perspectives. Within these subgroups, N^3^ quantifies normativity via local sample density estimation, determining how frequently similar value combinations occur within local neighborhoods of a measurement space, thereby capturing multiple co-existing normative constellations and yielding a continuous, graded measure of normativity.

#### 2.1.1 Local density estimation in subgroups

The algorithm quantifies the local sample density in different subgroups, defined as submatrixes *X*_*c*_ ∈ ℝ^*n×m*^, *c* = 1, 2, …, *g* of a reference dataset *X* ∈ ℝ^*a×b*^. We normalize each submatrix *X*_*c*_ column-wise using min–max scaling so that all entries satisfy *X*_*cij*_ ∈ [0, 1] for *i* = 1, 2, …, *n, j* = 1, 2, …, *m*.

Within each subgroup, we use a Nearest Neighbor approach and define a subset *N* for each sample *q*, such that it contains the *k* points in *X*_*c*_ which are the closest to sample *q* [25, 26]. Distance *d* is measured using the Euclidean distance. We define the local sample density *λ*(*q, c*) of sample *q* in subgroup *c* as the inverse of the average distance to its *k* nearest neighbors.

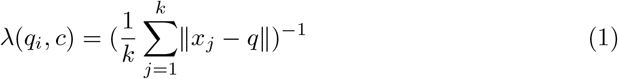

We calculate the local sample densities *λ* as described above for all samples in a subgroup *c* and get Λ_*c*_ = *{λ*(*q*_*i*_, *c*) | *i* = 1, 2, …, *n}*.

To ensure comparability between the different subgroups, we divide Λ_*c*_ by the median value.

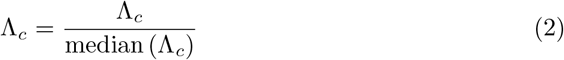

As a result we have a set of normalized local sample density estimations for *g* subgroups Λ′ = {Λ_*c*_ | *c* = 1, 2, …, *g*}.

To establish global context across the *g* subgroups, we aggregate the normalized local density estimates and model their empirical distribution using a parametric framework. Specifically, we fit the exponentiated Weibull distribution [27] to the values in Λ′, leveraging its flexibility to accommodate diverse distributional morphologies including skewness, heavy tails, and multimodality commonly observed in density-based statistics.

The probability density function of the exponentiated Weibull distribution is given by:

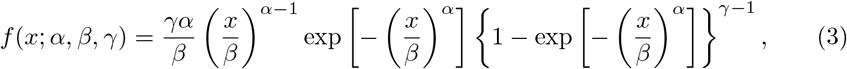

where *x* ∈ ℝ^+^ denotes the normalized local density estimate, *α >* 0 is the shape parameter governing distributional skewness, *β >* 0 is the scale parameter, and *γ >* 0 is the exponentiation parameter controlling tail behavior. We denote the complete parameter vector as *θ* = (*α, β, γ*)^⊤^ and estimate it via maximum likelihood estimation. Using the fitted distribution *f* (·; *θ*^∗^), we transform each normalized density estimate into a likelihood value, thereby establishing a common probabilistic scale across all subgroups and samples.

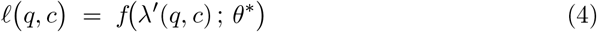

where *f* (·; *θ*) denotes the exponentiated Weibull density with parameters *θ, θ*^∗^ are the fitted parameters, and *ℓ*(*q, c*) is the likelihood assigned to the normalized local density of sample *q* in subgroup *c*.

To keep as much information as possible, we add a sign to *f*, which indicates in which direction a sample is deviating from the median. In this context, samples whose local sample density is smaller than the median, receive a negative value, while samples whose local sample density is larger than the median, have a positive value.

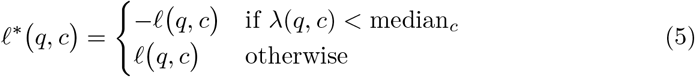

Finally, to foster intuitive interpretation, we scale the signed likelihood *ℓ* to an interval of [-1, 1]. The final value is a normativity estimation on how common the configuration of sample *q* appears within a particular control group *c*.

#### 2.1.2 Normativity Profile and N^3^ biomarker

In the next step, we aggregate normativity estimations from several subgroups into a vector, which we call normativity profile and which depicts an individual’s normativity measured from multiple meaningful angles or viewpoints.

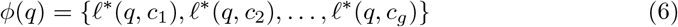

To synthesize the multidimensional information entailed in an individual normativity profile into a single, actionable metric, we repeat the local density estimation approach outlined above. In a second layer of normativity estimation, we use the normativity profile *ϕ* as input data and repeat the local sample density estimation approach outlined above. Now, the local density estimation is based on the *g* values of *ϕ*, computed in the normativity profile space. Thereby, we measure the commonness of a normativity profile in relation to other normativity profiles seen in a particular reference population. This can either be done globally (on all normativity profiles of the sample), or again in subgroups. The output of this meta-normativity estimation is the return value of the N^3^ algorithm, what we call the N^3^ biomarker.

#### 2.1.3 Training vs. Inference Phase

The N^3^ algorithm is trained using a normative reference sample. Using this sample, we learn scaling parameters, neighborhood structure, and distributional parameters, first in the original measurement space and second in the normativity profile space. Specifically, we first use the values in the reference sample to learn how frequently different medical configurations occur within each subgroup. We then aggregate the resulting values into a normativity profiles and fit a second layer of local density estimation on these profiles.

At inference, a new individual is projected into the learned spaces. We apply the subgroup definitions, the scaling parameters estimated during training and compute the local densities and signed likelihoods using the pre-fitted distributions. During this process, we first obtain a normativity profile, which is subsequently passed through the second-layer models, again using only parameters learned in the training phase, to yield the individual’s N^3^ biomarker value.

#### 2.1.4 Application to Brain Structure

In our application to brain structure, we stratify the training sample by sex and age, resulting into a total of 100 control groups containing same-aged females or males (22 to 72 years), respectively. Each sample is characterized by 5 different features, namely the brain structural volumes (GM, WM, WMH, CSF, TIV) of each individual. To mitigate different sample sizes of different age groups, we join either the lower, the upper, or both neighboring age groups of underrepresented age groups, so that the sample size per age group approximates the median sample size available per sex. We set the k parameter to 10% of the control group sample size, but limit its upper bound to 15 to prevent too broad comparisons *k* = *min*(round(0.1 × *n*), 15).

Applying the N^3^ algorithm, we then first evaluate the normativity of an individual’s brain features in comparison to the configurations seen in the continuum of age-groups of the same sex, before we analyze the resulting normativity profiles in relation to same-aged peers.

### 2.2 Materials

T1-weighted MRI scans data from six different studies were provided by the respective consortia. The training data for fitting models for the different biomarkers comprised 30,047 samples from the population-based NAKO cohort (for details please see A) [28–30]. We exclude age groups below 22 years and above 72 years due to small sample sizes (n *<* 100), which restricts the final sample to 29,883.

Using T1-weighted MRI scans from 7,013 additional individuals from different study populations, we validate each biomarker’s ability to differentiate between healthy inter-individual variability and pathological norm deviations. To this end, instances of Mild Cognitive Impairment (MCI), Alzheimer’s Disease (AD) and Frontotemporal Dementia (FTD) serve as model diseases to represent brain structural alterations and pathological norm deviations. Data was sourced from the Alzheimer’s Disease Neuroimaging Initiative (ADNI) [31], the Australian Imaging, Biomarker & Lifestyle Study of Aging (AIBL) [32], the Frontotemporal Lobar Degeneration Neuroimaging Initiative (NIFD), and the Open Access Series of Imaging Studies 3 (OASIS3) [33, 34] (for details see section A and Table A1). In general, if more than one measurement was available per participant, we restrict each study’s dataset to the first (baseline) measurement of the participant. Exclusion criteria were applied based on age; participants younger than 22 years or older than 72 were omitted from the study, due to insufficient sample sizes in the normative reference sample.

To evaluate the robustness of the N^3^ brain structural normativity assessments, we use artificially downsampled subgroups of the NAKO study for training. Validation subsets included n=835 healthy control participants from the the Münster-Marburg Affective Disorder Cohort (MACS) [35] study which predominantly comprises younger and middle-aged adults, and an additional n=1073 healthy older adults from the ADNI study to span a wider age demographic (see Methods section 2.5).

#### 2.2.1 Image Preprocessing

All images underwent preprocessing using the standard software CAT12 (version: cjp v0008, spm12 build v7771; cat12 build r1720) with default parameters. In short, images were bias-corrected, tissue classified, and normalized to MNI-space using linear and non-linear transformations. In this work, we use volume metrics of gray matter (GM), white matter (WM), white matter hyperintensities (WMH), total intracranial volume (TIV) and cerebrospinal fluid (CSF) derived from the SPM12-based tissue segmentation procedure.

### 2.3 Brain Age Model

In the Brain Age paradigm, the brain structure is evaluated with respect to aging effects seen in a healthy reference sample. This is realized by means of a machine learning model trained to predict chronological age from brain structure. The deviation between chronological and predicted age is referred to as the Brain Age Gap (BAG). While a small BAG is considered normative and age-appropriate, a larger positive or negative BAG symbolizes premature or delayed neurostructural degeneration, respectively. The resulting normativity estimation, i.e. the BAG values, have been associated with numerous neurological and psychiatric conditions [15, 16]. For comparison with N^3^, we train a Brain Age Model using the Python library photonai [36]. We use a Support Vector Machine (SVM), for which we optimize the C and gamma parameters in the nested-cross-validation procedure (k=10 outer folds and two randomly shuffled inner folds with a test size of 0.1). The best model achieves an average MAE of 5.43. We use age-stratified 90% of the available normative dataset for model training. We use the remaining 10% of the normative dataset to train a linear age bias correction as described in Peng et al. [37]. For the evaluation of unseen samples, we use the Brain Age SVM model to predict age and subsequently apply the bias correction model, before we calculate the difference between the chronological and predicted age, the BAG.

### 2.4 Normative Modeling

We calculate normative models on the training data using the Predictive Clinical Neuroscience toolkit as described in Rutherford et al. [10]. To train the models, we normalize GM, WM, WMH, CSF by Total Intracranial Volume (TIV) and fit Bayesian Linear Regression models with default parameters. Subsequently, we derive z-scores for each variable and aggregate them into four normative modeling markers, each yielding a single score per individual. The best performing marker counts the number of absolute z-scores greater than 1.96, reflecting the burden of outlying features per person, and is discussed throughout the manuscript. We evaluated three more aggregation strategies. The second marker sums the absolute z-scores across all features, capturing the overall magnitude of deviation from the normative reference. The third marker is defined as the average z-score, summarizing the mean direction and extent of deviation. Finally, we obtain a low-dimensional embedding of the multivariate z-score patterns using a UMAP component, which serves as a fourth, non-linear aggregation of normative deviations. For completeness, Table C4 and C5 in Appendix C reports the statistical and machine-learning performance of all evaluated markers.

### 2.5 Statistical Analysis

A Type III Sum of Squares ANOVA was performed using an ordinary least squares model to assess the discriminative and explanatory power of each normativity marker in distinguishing patients from controls. The model was adjusted for potential confounders, including age, age squared (to mitigate non-linear effects), sex and scanner. Partial eta squared (*η*^2^) was used to quantify effect size, providing an estimate of how much variance in disease progression could be explained by each normativity marker, alongside a 95% confidence interval.

We evaluate and rank the different normativity markers by post-hoc comparisons of their effect size. To test the observed marker differences for statistical significance, we calculate the ANOVA for each marker with 1000 random permutations. To determine the p value of the marker differences, we evaluate the actual difference between the *η*^2^ of our marker N^3^ and the *η*^2^ another marker, with those found in the 1000 random permutations.

To assess each normativity marker’s consistency across age groups, an analysis of age bias was conducted using Spearman’s rank correlation to evaluate the correlation between the normativity estimation values and age in healthy controls.

To assess stability of the N^3^ models, the Intraclass Correlation Coefficient (ICC) model (2,1) was applied. For this purpose, we used the NAKO sample to train the normativity models, which were downsampled to mimic smaller study populations. Particularly, we divide the training set in k=[10, 5, 3, 2] non-overlapping parts of equal size, train normativity models within each of these subsets, and use hold-out data to ensure the stability of the normativity estimates. To ensure validity of the test, we use only age groups with more than 500 samples available from the training sample and more than 20 samples in the test samples.

All statistical analyses were implemented in Python using the *scipy, statsmodels* and pingouin libraries.

### 2.6 Machine Learning Analysis

Machine learning models transcend conventional statistical models by handling multivariate and non-linear relationships and shifting the focus from group average comparisons to predictions on an individual level[38]. We employ cross-validation strategies, which systematically tests each marker against new, unseen data to verify the accuracy, robustness, and generalizability of the models [39]. Our analytical pipeline employed the open-source Python framework photonai [36]. The analysis involved nested cross-validation to robustly estimate model performance and avoid overfitting, using k=5 outer folds and k=10 inner folds, each fold stratified to entail a balanced proportion of samples from the diseased class. Hyperparameter optimization was performed via Grid Search to fine-tune the support vector machine (SVM) parameters C and gamma. The machine learning pipeline included steps for z-normalization and balanced sampling (random under-sampling techniques) to address class imbalance within the training data. Models were trained with each of the biomarkers as the only feature, enabling us to quantify each biomarker’s ability to capture disease signals of neurodegenerative diseases. We measure balanced accuracy, recall, precision and f1 score of each of the normativity markers in the classification of neurodegenerative diseases.

## 3 Results

### 3.1 The N^3^ biomarker for brain structure and its biological interpretation

In a first step, the N^3^ framework analyzes the frequency of different brain structural configurations within sex-matched subgroups of successive chronological age. The result is a vector that expresses how commonly an individual’s specific configuration occurs within each demographic subgroup, i.e. how their measurements are embedded within the natural variety seen along the aging continuum. We call this vector a normativity profile. Its shape indicates how an individual’s configuration is positioned relative to expected trajectories of brain structural decline. For example, an individual’s configuration may show similarity with those occurring frequently in younger age groups, indicating younger brain features and fewer aging effects as commonly seen in same-aged individual (see Fig. 2). However, normativity profiles that shift away from an individual’s chronological age are not necessarily indicative of disease. Since genetic and environmental influences on brain structure accumulate over time, we expect substantial variability in normativity profile shapes, particularly in older age groups. We therefore quantify how frequently different normativity profile shapes occur within each age group, thereby allowing for several normative variants of brain structural decline. In a second step, the N^3^ framework therefore assesses how common a given profile shape is within each sex- and age-specific subgroup. The resulting metric is the N^3^ biomarker, which provides an individualized index of how typical or atypical a person’s brain structural configuration appears relative to the variability observed in both neurophysiology and aging trajectories across individuals.

**Fig. 1.**
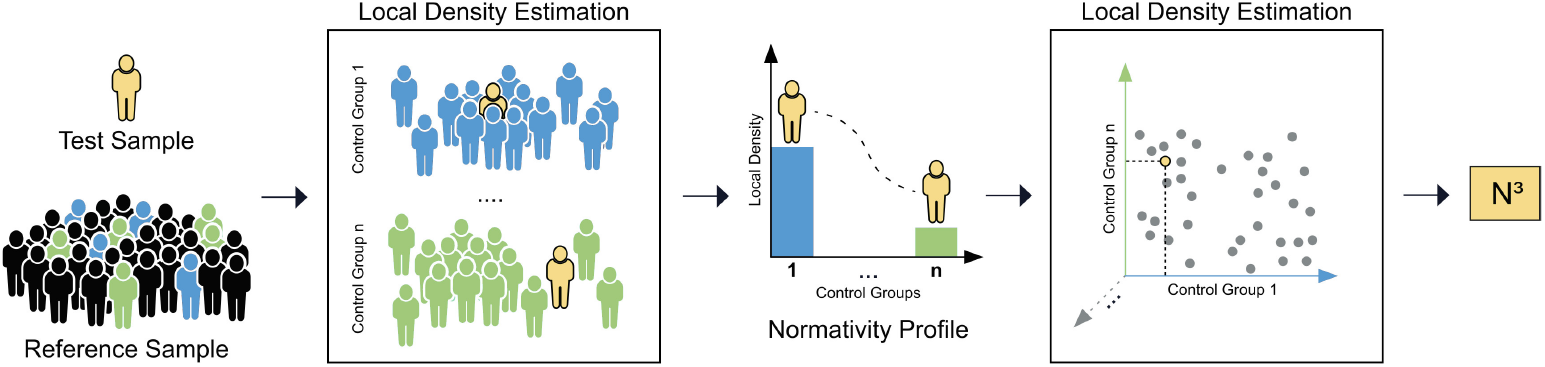
Schematic summary of the Nearest Neighbor Normativity N^3^ framework. The N^3^ framework analyzes the local sample density around individual measurements in strategically chosen subgroups of a normative reference dataset. This strategy accommodates multiple, equally viable health states in the population and promotes the detection of nuanced irregularities that may escape broader comparative models. Moreover, by evaluating the local sample density in multiple strategically chosen subgroups, the N^3^ framework provides a multifaceted contextualization of medical observations. We call the resulting vector a normativity profile. The information contained in the normativity profile is converted into a singular, actionable metric, by re-applying local density estimation. The resulting metric expresses how common a normativity profile is for a particular demographic or clinical subgroup. It can be adapted to a variety of clinical inquiries, such as how common is this profile amongst young women, treatment resistant patients or genetic predispositions.

**Fig. 2.**
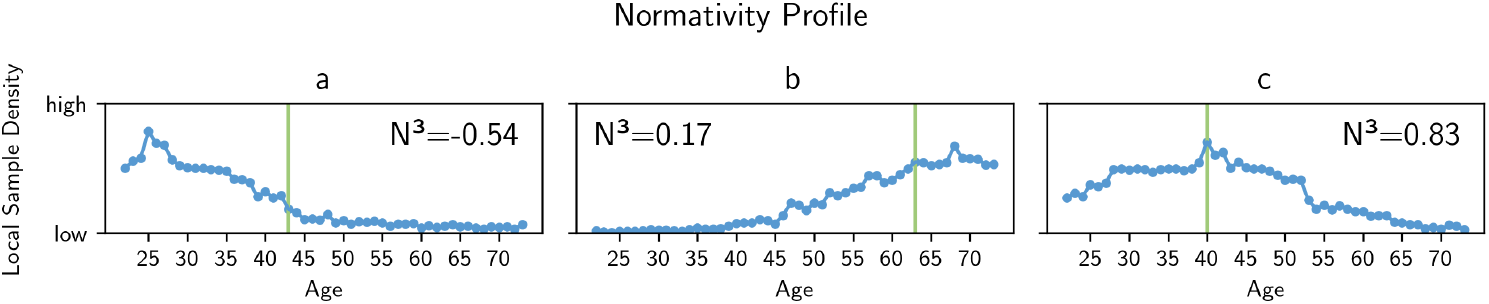
Exemplary normativity profiles of three individuals. For illustration, we show the normativity profiles of three individuals of the training sample. The N^3^ framework compares an individual’s brain structural features (global volumes of gray matter, white matter, white matter hyperintensities, cerebrospinal fluid and total intracranial volume) to those seen in different age groups along the age continuum. The normativity profile shows the local sample density in different age groups, i.e. how common similar volumetric configurations appear along the aging continuum (blue) as well as the individual’s chronological age (green). (**a**) An individual’s data shows higher local sample density in younger age groups, indicating that the volumetric configuration is more common within younger age groups and less common within same-aged individuals (**b**) An individual’s volumetric configuration appears more common within older age groups, indicating premature aging effects. (**c**) A brain structure appears most common within its own age group and deprecating occurrence within other age groups.

**Fig. 3.**
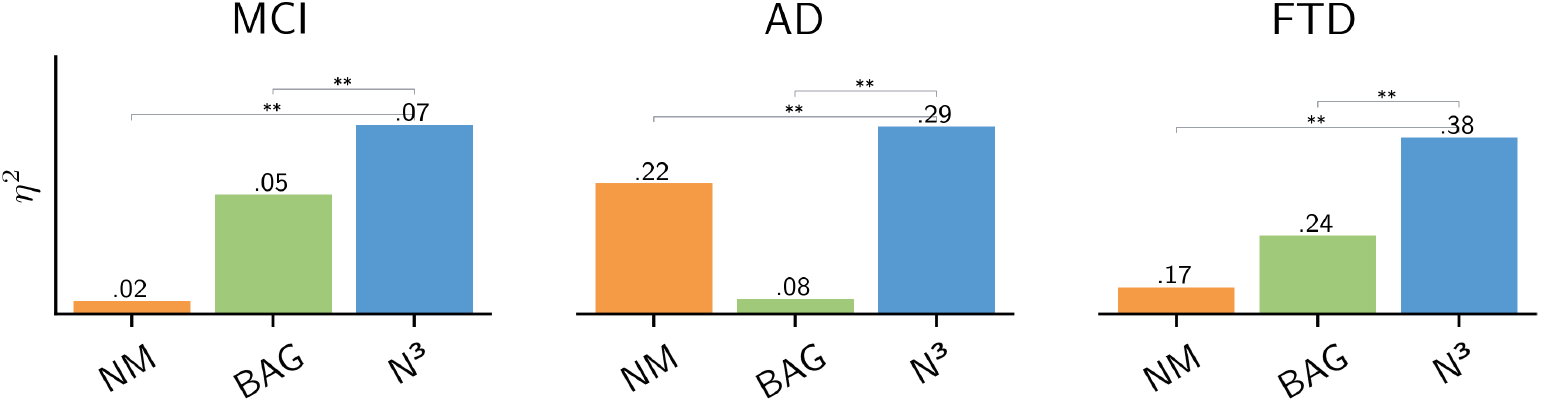
Results of the statistical analyses. Statistical effect sizes (partial eta squared - *η*^2^) are given for the different biomarkers (N^3^ - our approach, NM - Normative Modeling outlier score, and the BAG - Brain Age Gap). We evaluate each approach’s ability to differentiate between controls and diseased individuals using neurodegenerative diseases as representative models diseases, in particular Mild Cognitive Impairment (MCI), Alzheimer’s Disease (AD), and Frontotemporal Dementia (FTD). Post-hoc comparisons of the effect sizes revealed larger explained variance of our N^3^ marker in all neurodegenerative conditions. Significance in the differences was confirmed through permutation testing using 1000 random permutations and is indicated above (** *p <*.001).

### 3.2 Increased statistical explanatory power in distinguishing neurodegenerative diseases

For AD, the N^3^ marker showed the largest effect size (*η*^2^= 0.29), indicating that approximately 29% of the variability between the AD group and controls can be explained by differences in the N^3^ scores (see Figure 4 and Table 2). In the context of FTD, all markers demonstrated large effect sizes, while the N^3^ stood out with an effect size of *η*^2^ = 0.38. The results for MCI differed, as all markers showed generally low explanatory power. The N^3^ marker displayed a relative advantage, with an effect size of *η*^2^ = 0.07, compared to *η*^2^ = 0.05 for the BAG and *η*^2^ = 0.02 for the normative modeling score. Overall, the N^3^ biomarker showed the largest effect sizes across all examined neurodegenerative conditions.

**Fig. 4.**
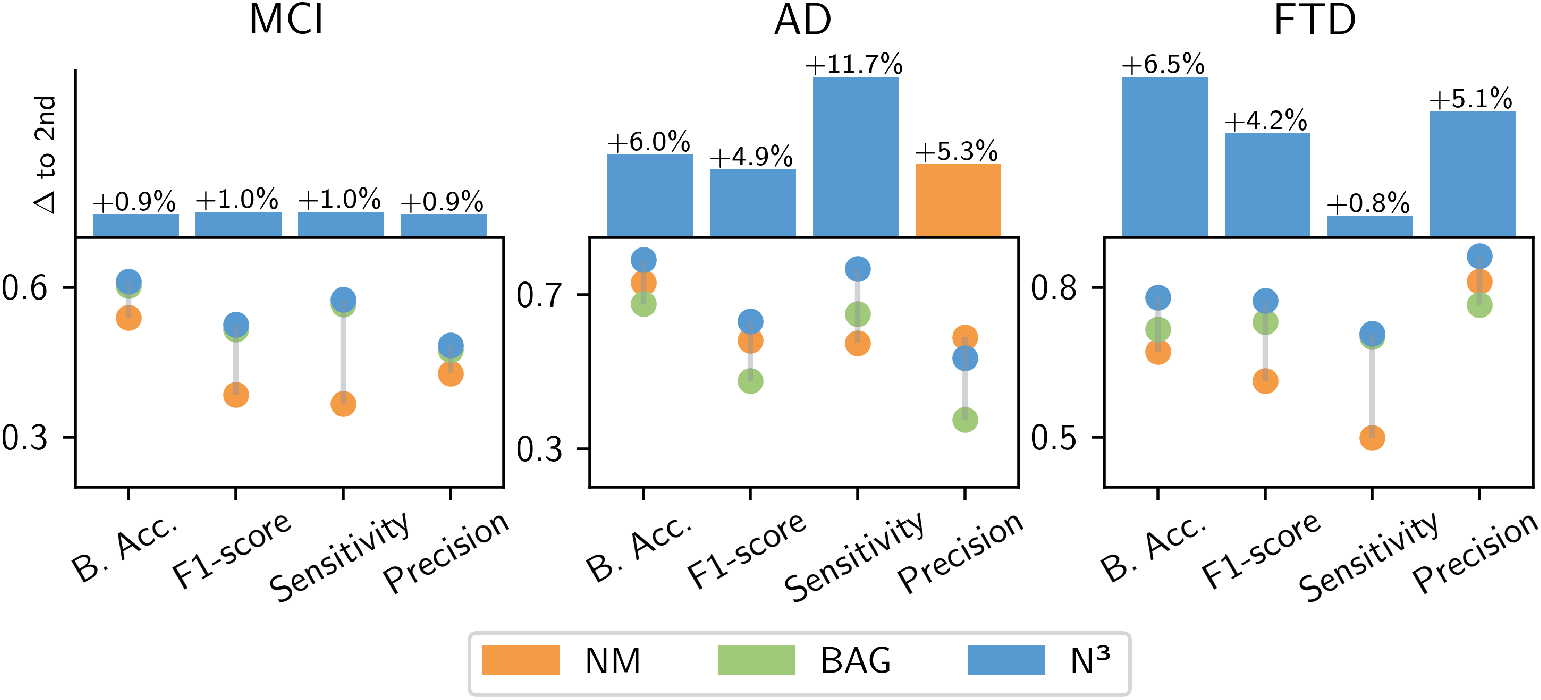
Results of machine learning analyses. We use machine learning to evaluate the expressiveness of each biomarker on a single-subject level (N^3^ - our approach, NM - Normative Modeling outlier score, and the BAG - Brain Age Gap). We evaluate each approach’s ability to differentiate between cognitively unimpaired and diseased individuals using neurodegenerative diseases as representative models diseases, in particular Mild Cognitive Impairment (MCI), Alzheimer’s Disease (AD), and Frontotemporal Dementia (FTD). The N^3^ marker demonstrated increased accuracy in predicting the occurrence of pathological norm deviations. We show the different marker’s performance metrics [balanced accuracy (B.Acc), F1-Score, Recall and Precision] and the performance advantage,.i.e. the delta, of the best biomarker in relation to the second best biomarker in percentage (Δ to 2nd).

**Table 1.**
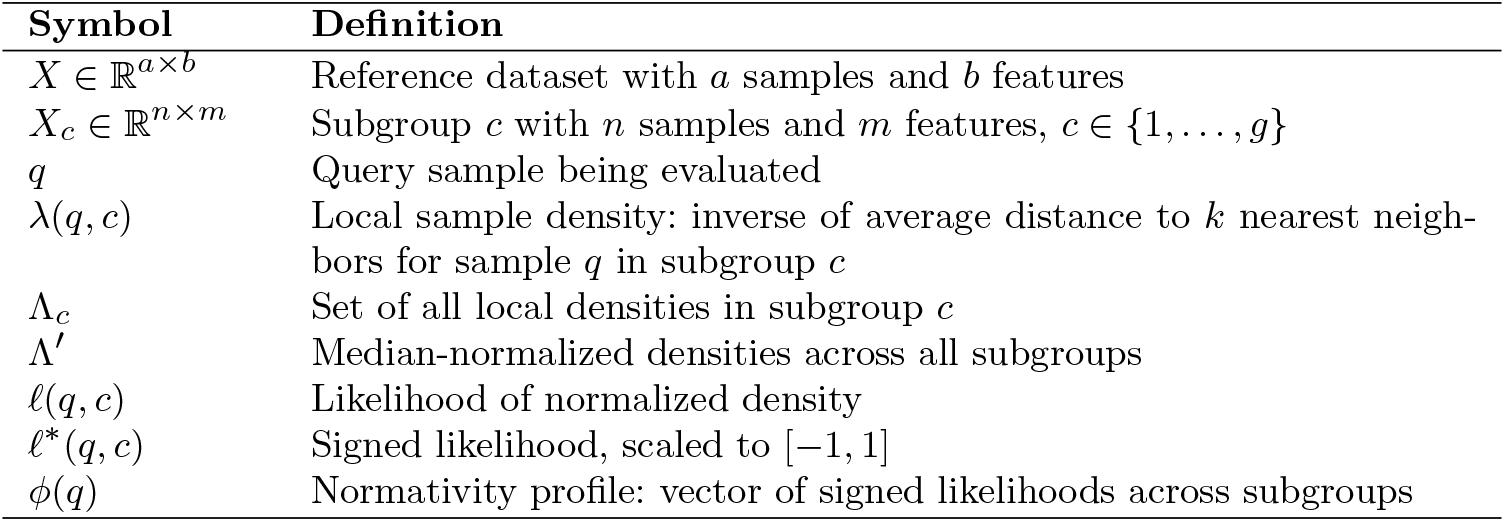
Notation and variable definitions.

**Table 2.** Statistical discrimination of neurodegenerative diseases. We evaluate the markers’ ability to distinguish inter-individual variability from pathological norm deviations in statistical analyses. We evaluate each approach’s ability to differentiate between controls and diseased individuals using neurodegenerative diseases as representative models diseases, in particular Mild Cognitive Impairment (MCI), Alzheimer’s Disease (AD), and Frontotemporal Dementia (FTD). We compare N^3^ - our approach, NM - Normative Modeling outlier score, and the BAG - Brain Age Gap, respectively. All F-statistics and effect sizes *η*^2^ are significant (p*<*0.001). Best performance is indicated in bold.

| Marker | MCI |  | AD |  | FTD |  |
| --- | --- | --- | --- | --- | --- | --- |
| | F | $\eta^2$ | F | $\eta^2$ | F | $\eta^2$ |
| NM | F(1,4565)=74 | 0.016 | F(1,3709)=1,073 | 0.223 | F(1,580)=121 | 0.173 |
| BAG | F(1,4565)=220 | 0.046 | F(1,3709)=328 | 0.081 | F(1,580)=184 | 0.242 |
| $N^3$ | F(1,4565)=**313** | **0.067** | F(1,3709)=**1,412** | **0.292** | F(1,580)=**346** | **0.376** |

### 3.3 Single-subject predictions

The efficacy of the N^3^ marker was confirmed at the individual level (see Figure 4 and Table 3). In the specific cases of AD and FTD, the N^3^ marker demonstrated improvements in balanced accuracy scores, surpassing the second-best markers by 6.0% for AD and 6.5% for FTD. For MCI the efficacy of all markers notably declined, in line with the small effect sizes observed in statistical analysis. Here, the N^3^ reached an 0.9% improvement to the next best marker, the BAG. With regard to the F1-scores, the N^3^ marker achieved the highest performance in all neurodegenerative diseases. N^3^’s precision for AD was 5.3% behind the normative modeling marker and it was superior by 5.1% for FTD and 0.9% in MCI compared to the second best result. With regard to sensitivity, the N^3^ marker displayed highest rates in AD (11.7%), and slightly improved rates in MCI and FTD (+1.0% and +0.8%). Overall, the N^3^ biomarker achieved the highest predictive performance in the classification of neurodegenerative diseases among the compared markers in our evaluations.

**Table 3.**
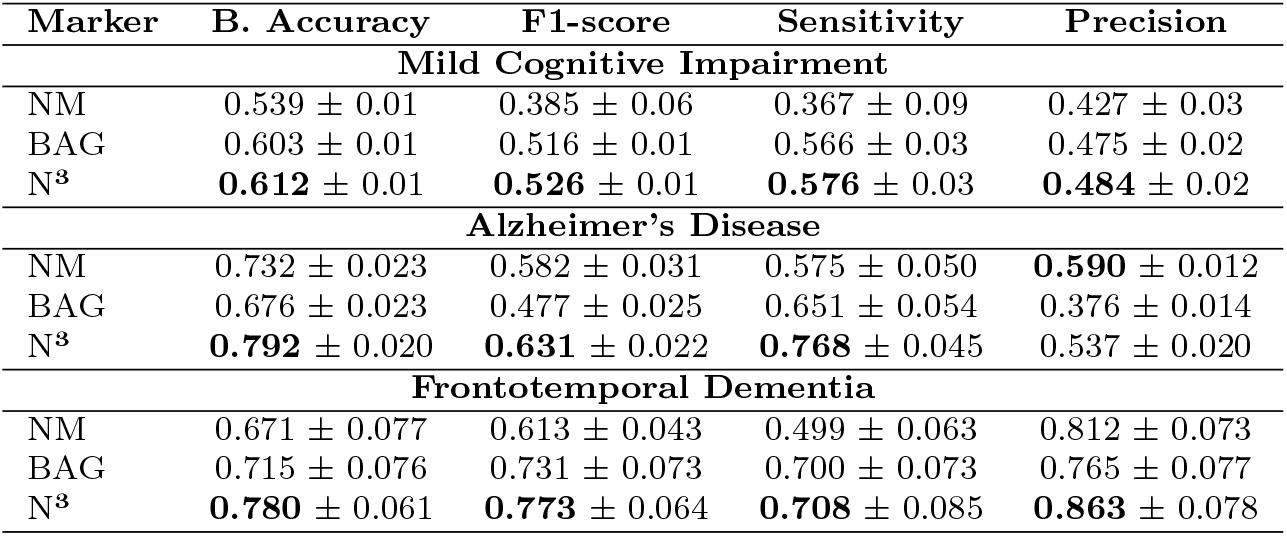
Performance metrics of the machine learning analyses. We train a machine learning classifier with each of the biomarkers as single feature to predict the occurrence of neurodegenerative diseases in individual cases. Highest performance is indicated in bold. We compare N^3^ - our approach, NM – Normative Modeling outlier score, and the BAG - Brain Age Gap, respectively. Best performance is indicated in bold.

| Marker | B. Accuracy | F1-score | Sensitivity | Precision |
| --- | --- | --- | --- | --- |
| **Mild Cognitive Impairment** |  |  |  |  |
| NM | 0.539 ± 0.01 | 0.385 ± 0.06 | 0.367 ± 0.09 | 0.427 ± 0.03 |
| BAG | 0.603 ± 0.01 | 0.516 ± 0.01 | 0.566 ± 0.03 | 0.475 ± 0.02 |
| N<sup>3</sup> | **0.612 ± 0.01** | **0.526 ± 0.01** | **0.576 ± 0.03** | **0.484 ± 0.02** |
| **Alzheimer’s Disease** |  |  |  |  |
| NM | 0.732 ± 0.023 | 0.582 ± 0.031 | 0.575 ± 0.050 | **0.590 ± 0.012** |
| BAG | 0.676 ± 0.023 | 0.477 ± 0.025 | 0.651 ± 0.054 | 0.376 ± 0.014 |
| N<sup>3</sup> | **0.792 ± 0.020** | **0.631 ± 0.022** | **0.768 ± 0.045** | 0.537 ± 0.020 |
| **Frontotemporal Dementia** |  |  |  |  |
| NM | 0.671 ± 0.077 | 0.613 ± 0.043 | 0.499 ± 0.063 | 0.812 ± 0.073 |
| BAG | 0.715 ± 0.076 | 0.731 ± 0.073 | 0.700 ± 0.073 | 0.765 ± 0.077 |
| N<sup>3</sup> | **0.780 ± 0.061** | **0.773 ± 0.064** | **0.708 ± 0.085** | **0.863 ± 0.078** |

### 3.4 Stability and Robustness

The calculation of the N^3^ marker relies on local density estimation. As such it is highly dependent on the composition of the reference sample. Therefore, we investigate how changes to the sample composition and sample size affect the stability of the N^3^ model. We retrained N^3^ models with downsampled subsets of varying size, thereby mimicking smaller studies and different study participants. We then apply the different normativity models and predict normativity on an external dataset. Particularly, we evaluate if predictions remain consistent across different sample sizes and sample compositions. We quantify the stability of the normativity estimates by calculating the Intraclass Correlation Coefficient (ICC) (see Methods 2.5). Results are visualized in Figure 5b. We see that random samples of 200 individuals and above show consistently high stability (ICC of 0.75 and above). Moreover, the ICC approaches 0.9 in larger sample sizes, starting at 150 participants. While the results are calculated for the use case of brain structural normativity estimation, they are a first indication density-estimation based normative models can be realized by dividing larger samples into subgroups of a few hundred samples and above.

**Fig. 5.**
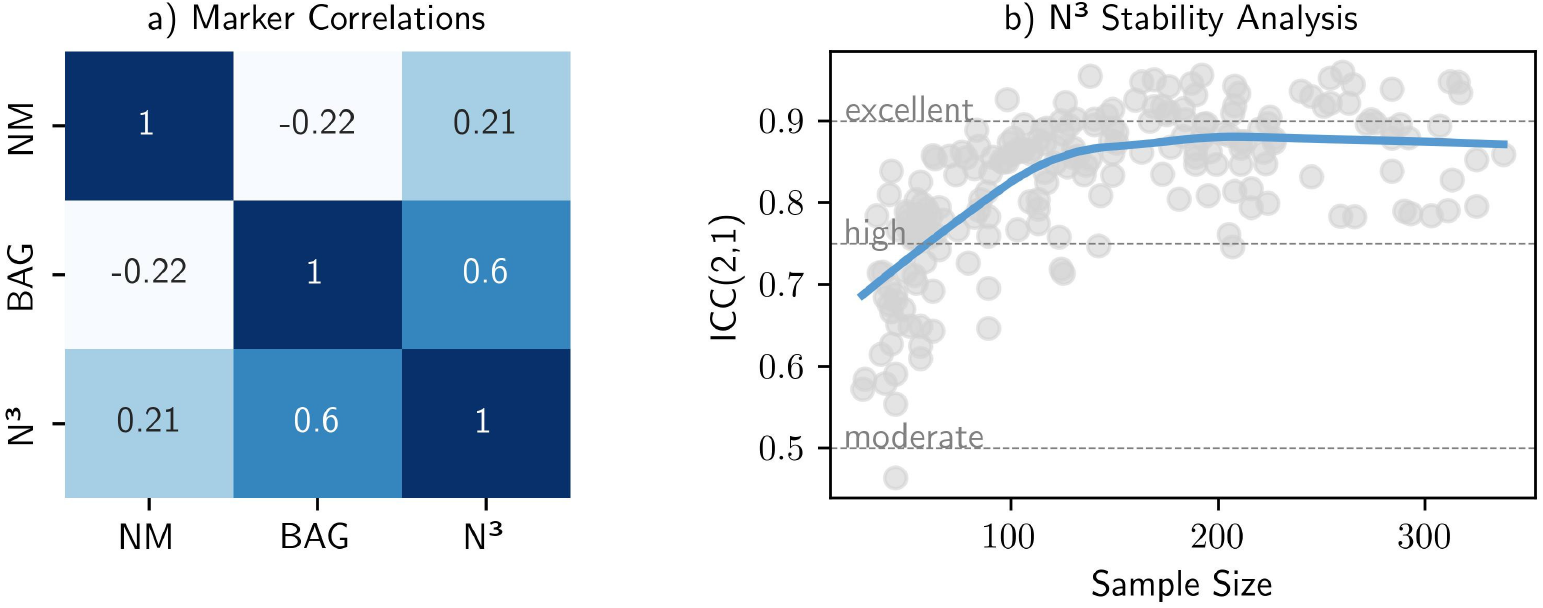
Robustness and consistency of the N^3^ framework. (**a**) We calculated the correlation matrix among the different biomarkers, which emphasize the distinctiveness and complementarity of the different markers. (**b**) We tested the impact of sample size and sample composition on the reliability of the N^3^ biomarker by calculating the intraclass correlation coefficients across repeatedly downsampled subsets of the training data. We see that the N^3^ marker exhibits an ICC ≥ 0.75 for n*>*100 individuals and approaches ICC=0.9 for n*>*150 individuals.

In terms of inter-marker relationships (Figure 5a), the correlation analysis revealed generally weak associations between the normative modeling marker and the other markers (| *ρ* | ≈ 0.20), indicating that these indices capture largely distinct aspects of brain structural norm deviations. By contrast, the BAG and the N^3^ marker show a moderate positive correlation (*ρ* = 0.6), suggesting that N^3^ partly aligns with established notions of accelerated brain aging while still retaining unique variance. Taken together, these patterns point to a set of complementary biomarkers with overlapping but non-redundant information content, supporting the idea that combining N^3^ with existing markers such as BAG and traditional normative indices may yield a richer, more nuanced characterization of brain structural alterations.

Finally, it is essential for normativity estimations to remain consistent across different age groups and avoid age biases that could complicate semantic interpretations. An analysis of the age correlation of the N^3^ marker indicates its stability over the age range, showing no significant association to age (*ρ*=0.009, p=0.60). In comparison, the normative model marker shows a significant but smaller correlation to age (*ρ*=0.104, p*<*0.001), while the Brain Age Gap (BAG) exhibits a moderate age bias (*ρ*=-0.21, p*<*0.001).

Overall, these analyses show that the N^3^ biomarker yields stable normativity estimates for moderate sample sizes per age group, while remaining weakly to moderately correlated with other markers and thereby capturing partially distinct aspects of brain structural variation.

## 4 Discussion

We have introduced the N^3^ framework, which is designed to parse the natural heterogeneity in medical observations across individuals into diversity-aware normativity assessments. We applied it to brain structure, which resulted in an informative biomarker assessing the normativity of neurophysiological measurements. The resulting N^3^ biomarker for brain structure enabled superior differentiation between natural inter-individual variability and neurodegenerative diseases.

The N^3^ framework is based on two key algorithmic principles.

First, it relies on strategic contextualization, meaning that medical measurements are compared from multiple meaningful viewpoints within intentionally chosen subgroups of a reference dataset. This promotes the detection of nuanced particularities that may escape broader comparative models and provides a multifaceted positioning of medical observations, elucidating different aspects of normativity. The resulting normativity profile can, in turn, be interrogated for a wide range of clinically relevant questions, such as how frequently a given profile occurs among young women, in treatment-resistant patients, or in individuals with specific comorbidity patterns.

Second, N^3^ assesses the normativity of medical measurements using local sample density estimation techniques. By examining how often similar value combinations occur in small neighborhoods across the measurement space, it can flexibly represent multiple co-existing normative constellations while providing a flexible and continuous measure of normativity. Based on a continuum of natural variants in physiological organization, deviations can thus be quantified with greater granularity compared to traditional approaches.

In the application of the N^3^ framework to brain structure, we chose demographic subgroups as the primary context for interpretation. In this setting, the algorithm quantifies how common a particular combination of global brain volumes is across the diversity of brain structural measurements observed among same-sexed individuals along the aging continuum. The resulting vector, which we refer to as the normativity profile, encodes the resemblance of an individual’s brain to different natural atrophy progression states for that sex across the lifespan. In the second step, we contextualize each normativity profile against those of same-aged individuals to determine how frequently a given profile shape occurs within that age group. The resulting biomarker quantifies the typicality of an individual’s aging pattern among their peers. In clinical applications, the same procedure could be adapted to alternative contextualizations, such as profiles typical for specific comorbidity constellations or genetic or environmental risks.

Because N^3^ relies on local density estimation, it can represent similar aging patterns in neighboring age groups without forcing a trade-off in normativity metrics between them. More specifically, a particular configuration can be equally typical in several adjacent age groups. This contrasts with brain age models, which must assign a single best-fitting age and compress the richness of aging trajectories into a point estimate. Moreover, the N^3^ framework is able to detect irregular configurations that are uncommon across all age groups, and differentiate between general irregular anatomy and age-inappropriate configurations. Finally, the N^3^ framework is through its conceptualization able to revealing potential shifts towards constrained and pathologically similar configurations, if disease erodes the natural spectrum of variability and converts uniqueness into uniform manifestations.

In general, the interpretation and contextualization of individual brain structures holds significant potential for various domains. As stated above, a reliable biomarker for brain structural normativity is eagerly sought in neuropsychiatric research. Here, biomarkers may enable comprehensive assessments of neurostructural alterations associated with specific symptoms, to better understand the etiology and pathogenesis of different disease phenotypes [12, 38, 40]. For example, a valid and robust neurostructural biomarker would facilitate the analysis of environmental influences, genetic predisposition or neuroinflammatory contributions to understand disease mechanics and optimize individual disease management strategies [16, 23, 41, 42].

In the realm of neurodegenerative diseases, the ability to detect brain structural alterations early is of critical clinical relevance, as it has been shown that structural changes in the brain can manifest well before clinical symptoms become apparent [43, 44]. Furthermore, evidence supports the presence of multiple underlying neuropathological processes [21, 45], underscoring the methodological importance for models accommodating multiple disease prototypes. Here, a reliable brain structural screening tool could be attached to routine MRI scans and support disease interception to promote the prevention or delay of disease progression [46–49].

Our approach accommodates the multivariate nature of brain structures [50] and aligns with other modern understandings of heterogeneity, such as the concept of neurotypicality [51–53]. Traditionally seen as a uniform standard, brain architectures are now understood to encompass a spectrum of neurological function and structures, reflecting the rich diversity of the human nature. Moreover, our findings resonate with recent work by Yang et al., where the authors found a range of multiple, co-occurring patterns of brain aging [54]. Their research underscores the significant inter-individual and also intra-individual variability, underscoring the complexity and uniqueness of individual neurodegenerative processes beyond population averages.

In this initial proof-of-concept study, we used a multi-site, large-scale population cohort as a reference. While it is recruited to be representative, it is a national cohort and cannot reflect the entire spectrum of biological diversity. Future work should extend this approach to multi-cohort and internationally inclusive reference datasets in order to broaden generalizability and fairness across different populations. Moreover, to advance translation to clinical practice, it is essential to evaluate how effectively normative reference models derived from study cohorts can interpret data from clinical care routine.

In general, our framework is constrained by the availability of sufficiently large reference samples to reliably estimate the frequency of different multivariate value configurations within subgroups. While large-scale population studies have become increasingly accessible, clinical investigations are often necessarily conducted in smaller, more deeply phenotyped cohorts, where resource limitations restrict sample size. In such settings, smaller studies inherently depend on the existence of a larger, demographically and clinically appropriate reference cohort to enable meaningful normative deviation estimates. In our evaluations, the N^3^ local density estimation algorithm exhibited high stability in samples of a hundred individuals, indicating substantial robustness in moderately sized research cohorts. Moreover, our framework can partially offset the limitations of smaller clinical samples by providing a principled way to interpret individual- and subgroup-level patterns relative to a broader population distribution, thereby enhancing the inferential value of these studies.

Notably, our evaluations are based on only five variables reflecting global brain structure volumes. As such, they represent the character of many clinical measurements which are broad aggregates of more complex physiological features. In our evaluation, the N^3^ approach has demonstrated its ability to effectively decode the information contained in these limited neurophysiological variables and was able to extract more meaningful insights compared to conventional approaches. However, in future work, the N^3^ framework needs to replicate these results for example in the processing of high-dimensional data. To this end, deep-learning based image representation could be used to evaluate the efficacy of the N^3^ framework in parsing whole-brain, 3D images. Moreover, to further evaluate and advance the clinical utility of the resulting N^3^ biomarker, its relation to genetic, environmental and other clinical risk factors [54–57] needs to be robustly invested. Finally, the current implementation of N^3^ limits spatial interpretability regarding the anatomical locus of detected abnormalities. Future extensions should incorporate regional granularity through region-of-interest (ROI) based analyses, applying the N^3^ framework to parcellated brain volumes or adding ROIs as an additional stratification dimension, thereby capturing region-specific aging trajectories and spatial vulnerability patterns. Generally, adding model-agnostic interpretability methods such as Shapley values would transform N^3^ into a feature-resolved normativity mapping framework.

Finally, the effectiveness of the N^3^ framework is dependent on the choice and robustness of the local density estimation strategy. In our application, the Nearest Neighbor Algorithm depends on the *k* parameter, which defines the number of neighbors considered in the quantification. Limiting the number of neighbors to 10% with an upper bound to 15 prevented overly broad comparisons while maintaining sufficient robustness across all control groups. Sensitivity analysis demonstrates robust N^3^ performance across several variations in *k* values, with increasing *k* systematically trading precision for recall (see Section B in Appendix). When applied to novel scenarios, the underlying algorithm should be customized or adapted to the unique requirements, e.g., by optimizing the nearest neighbor parameter *k*, choosing other local density estimation algorithms, using custom distance metrics or leveraging dimensionality reduction techniques [26, 58].

Conceptually, our work advances the field by redefining normativity as a landscape of physiologically permissible configurations rather than proximity to a single group-average prototype. By demonstrating that such diversity-aware normativity concepts can be translated into an effective biomarker of brain structure, our results provide a concrete pathway for moving from population-level averages toward individualized, configuration-based characterization of neurodegenerative and potentially other medical conditions.

## 5 Conclusion

The insights presented in this work show the potential of reevaluating how population norms are derived, applied, and interpreted in clinical practice. With our N^3^ framework, we operationalize normativity as a multidimensional, continuous landscape that emerges from the diverse configurations observed in a reference population. Applied to brain structure, the N^3^ biomarker was able to distinguish biologically coherent variants from truly atypical constellations and improved the detection of neurodegenerative diseases both in statistical as in well as in single-subject predictions. As we continue to refine and validate our N^3^ framework, it is our hope that the insights and ideas presented here will contribute to more diversity-aware normativity assessments and support our collective scientific endeavors to realize truly individualized patient care.

## Acknowledgements

This work was funded by the German Research Foundation (DFG, grant HA7070/2-2, HA7070/3, and HA7070/4 to T.H. and FOR2107 DA1151/5-1, DA1151/5-2, DA1151/9-1, DA1151/10-1, DA1151/11-1 to U.D.; SFB/TRR 393, project grant no 521379614), the Interdisciplinary Center for Clinical Research (IZKF) of the medical faculty of Münster (grant Dan3/022/22 to U.D. and MzH 3/020/20 to T.H.), and the IMF research instrument of the medical faculty of Münster (grant LE 1 1 24 09 to R.Leenings). X. Jiang was supported by the Deutsche Forschungsgemeinschaft (DFG) under Grant CRC 1450—431460824.

This project was conducted with data from the German National Cohort (NAKO) (www.nako.de). The NAKO is funded by the Federal Ministry of Education and Research (BMBF) [project funding reference numbers: 01ER1301A/B/C, 01ER1511D, 01ER1801A/B/C/D and 01ER2301A/B/C], federal states of Germany and the Helmholtz Association, the participating universities and the institutes of the Leibniz Association. We thank all participants who took part in the NAKO study and the staff of this research initiative.

Data used in the preparation of this article were obtained from the Alzheimer’s Disease Neuroimaging Initiative (ADNI) database (adni.loni.usc.edu). The ADNI was launched in 2003 as a public-private partnership, led by Principal Investigator Michael W. Weiner, MD. The primary goal of ADNI has been to test whether serial magnetic resonance imaging (MRI), positron emission tomography (PET), other biological markers, and clinical and neuropsychological assessment can be combined to measure the progression of mild cognitive impairment (MCI) and early Alzheimer’s disease (AD). Data collection and sharing for this project was funded by the Alzheimer’s Disease Neuroimaging Initiative (ADNI) (National Institutes of Health Grant U01 AG024904) and DOD ADNI (Department of Defense award number W81XWH-12-2-0012). ADNI is funded by the National Institute on Aging, the National Institute of Biomedical Imaging and Bioengineering, and through generous contributions from the following: AbbVie, Alzheimer’s Association; Alzheimer’s Drug Discovery Foundation; Araclon Biotech; BioClinica, Inc.; Biogen; Bristol-Myers Squibb Company; CereSpir, Inc.; Cogstate; Eisai Inc.; Elan Pharmaceuticals, Inc.; Eli Lilly and Company; EuroImmun; F. Hoffmann-La Roche Ltd and its affiliated company Genentech, Inc.; Fujirebio; GE Healthcare; IXICO Ltd.; Janssen Alzheimer Immunotherapy Research Development, LLC.; Johnson Johnson Pharmaceutical Research Development LLC.; Lumosity; Lundbeck; Merck Co., Inc.; Meso Scale Diagnostics, LLC.; NeuroRx Research; Neurotrack Technologies; Novartis Pharmaceuticals Corporation; Pfizer Inc.; Piramal Imaging; Servier; Takeda Pharmaceutical Company; and Transition Therapeutics. The Canadian Institutes of Health Research is providing funds to support ADNI clinical sites in Canada. Private sector contributions are facilitated by the Foundation for the National Institutes of Health (www.fnih.org). The grantee organization is the Northern California Institute for Research and Education, and the study is coordinated by the Alzheimer’s Therapeutic Research Institute at the University of Southern California. ADNI data are disseminated by the Laboratory for Neuro Imaging at the University of Southern California.

Data was in part provided by OASIS-3: Principal Investigators: T. Benzinger, D. Marcus, J. Morris; NIH P50AG00561, P30NS09857781, P01AG026276, P01AG003991, R01AG043434, UL1TR000448, R01EB009352. AV-45 doses were provided by Avid Radiopharmaceuticals, a wholly owned subsidiary of Eli Lilly.

Data used in preparation of this article were obtained from the Frontotemporal Lobar Degeneration Neuroimaging Initiative (FTLDNI) database. The investigators at NIFD/FTLDNI contributed to the design and implementation of FTLDNI and/or provided data, but did not participate in analysis or writing of this report (unless otherwise listed). Data collection and sharing for this project was funded by the Frontotemporal Lobar Degeneration Neuroimaging Initiative (National Institutes of Health Grant R01 AG032306). The study is coordinated through the University of California, San Francisco, Memory and Aging Center. FTLDNI data are disseminated by the Laboratory for Neuro Imaging at the University of Southern California The investigators at NIFD/FTLDNI contributed to the design and implementation of FTLDNI and/or provided data, but did not participate in analysis or writing of this report (unless otherwise listed). The FTLDNI investigators included the following individuals: Howard Rosen; University of California, San Francisco (PI) Bradford C. Dickerson; Harvard Medical School and Massachusetts General Hospital Kimoko Domoto-Reilly; University of Washington School of Medicine David Knopman; Mayo Clinic, Rochester Bradley F. Boeve; Mayo Clinic Rochester Adam L. Boxer; University of California, San Francisco John Kornak; University of California, San Francisco Bruce L. Miller; University of California, San Francisco William W. Seeley; University of California, San Francisco Maria-Luisa Gorno-Tempini; University of California, San Francisco Scott McGinnis; University of California, San Francisco Maria Luisa Mandelli; University of California, San Francisco

Data used in the preparation of this article was obtained from the Australian Imaging Biomarkers and Lifestyle flagship study of ageing (AIBL) funded by the Commonwealth Scientific and Industrial Research Organisation (CSIRO) which was made available at the ADNI database (www.loni.usc.edu/ADNI). The AIBL researchers contributed data but did not participate in analysis or writing of this report. AIBL researchers are listed at www.aibl.csiro.au.

## Declarations

### Data Availability

Data were obtained from the German National Cohort (NAKO), the Alzheimer’s Disease Neuroimaging Initiative (ADNI), the Open Access Series of Imaging Studies 3 (OASIS3), the Frontotemporal Lobar Degeneration Neuroimaging Initiative (NIFD, and the Australian Imaging, Biomarker Lifestyle Study of Aging (AIBL). Data of the MACS study are not publicly available. All other data are available upon request via the access management systems of the respective studies. (NAKO: nako.de/forschung, ADNI, AIBL, NIFD: ida.loni.usc.edu, OASIS3: sites.wustl.edu/oasisbrains)

### Code Availability

Code to realize the normativity estimation calculations within the N^3^ framework is written in the Python programming language and is provided as an open-source resource to the scientific community on Github (https://github.com/RLeenings/n3_framework.git).

## Appendix A Study Populations

### German National Cohort (NAKO)

The German National Cohort is a population-based longitudinal study initiated in 2014 aiming to investigate the risk factors for major chronic diseases in 200,000 persons living in Germany. It contains high-quality neuroimaging data from participants spanning a broad age range. In this study, we utilize the participants’ 3.0-Tesla T1w-MPRAGE MRI scans (voxel size 1×1×1 mm3, repetition time/ echo time=2300/2.98, flip angle=9°) [28–30].

### Alzheimer’s Disease Neuroimaging Initiative (ADNI)

ADNI is a major multicenter study started in 2003, designed to develop clinical, imaging, genetic, and biochemical biomarkers for the early detection and tracking of Alzheimer’s disease. The ADNI was launched as a public-private partnership, led by Principal Investigator Michael W. Weiner, MD. The primary goal of ADNI has been to test whether serial MRI, positron emission tomography (PET), other biological markers, and clinical and neuropsychological assessment can be combined to measure the progression of neurodegeneration. We included 1.5 and 3.0-Tesla T1w-MPRAGE MRI scans adhering to the ADNI sequence protocol, for scanner specific details please see https://adni.loni.usc.edu/data-samples/adni-data/neuroimaging/mri/mri-scanner-protocols/)

### Australian Imaging, Biomarker & Lifestyle Study of Aging (AIBL)

AIBL is an Australian study launched in 2006 focusing on understanding the pathways to Alzheimer’s disease. The cohort includes participants diagnosed with Alzheimer’s disease, mild cognitive impairment, and cognitively unimpaired elderly participants, providing insights into the aging process and the development of neurodegenerative diseases. AIBL study methodology has been reported previously [59]. MRI scans were performed using a 3D MPRAGE image (voxel size 1.2×1×1 mm3, repetition time/echo time=2300/ 2.98, flip angle=8°)[32].

### NIFD Dataset

The Frontotemporal Lobar Degeneration Neuroimaging Initiative (FTLDNI) was funded through the National Institute of Aging, and started in 2010. The primary goals of FTLDNI were to identify neuroimaging modalities and methods of analysis for tracking frontotemporal lobar degeneration (FTLD) and to assess the value of imaging versus other biomarkers in diagnostic roles. The Principal Investigator of NIFD was Dr. Howard Rosen, MD at the University of California, San Francisco. We use the provided 3D MPRAGE T1-weighted images (voxel size 1×1×1 mm3, repetition time/echo time=2300/2.9, matrix = 240 × 256 × 160) The data are the result of collaborative efforts at three sites in North America. For up-to-date information on participation and protocol, please visit http://memory.ucsf.edu/research/studies/nifd

### Open Access Series of Imaging Studies 3 (OASIS3)

OASIS3 serves as a comprehensive digital repository for MRI brain data that supports longitudinal studies of normal aging and cognitive decline [33, 34]. The project is distinguished by its wide age range of participants, providing diverse datasets that enhance the understanding of late-life brain diseases alongside physiological aging processes. We include 3D MPRAGE T1-weighted images (voxel size 1.0 or 1.2×1×1 mm3, repetition time/echo time=2300/2.95 or 2400/3.16 (depending on the scanner), flip angle=9°, FoV=240 or 256mm)

### Marburg-Münster Affective Disorder Cohort Study (MACS)

The MACS cohort is part of the DFG-funded research group FOR2107 cohort, researching the etiology and progression of affective disorders [35]. The goal is to integrate and understand the clinical and neurobiological effects of genetisc and environmental factors, and their complex interactions. Participants received financial compensation and gave written informed consent. We use the T1-weighted neuroimaging scans of n=835 healthy control participants to evaluate stability of the N^3^ models. Images were in Marburg (MR) or Münster (MS) (voxel size 1×1×1 mm3, repetition time/echo time=MR: 1900, MS: 2130/MR: 2.26, MS: 2.28, flip angle=8°, FoV = 256 mm, matrix = 256 × 256, slice thickness = 1 mm)

**Table A1.** Study Data Summary. T1-weighted MRI scans from six different studies were provided by the respective consortia. If more than one measurement was available per participant, we restrict each study’s dataset to the first (baseline) measurement of the participant. Exclusion criteria were applied based on age; participants younger than 22 years or older than 72 were omitted from the study, due to insufficient sample sizes in the normative reference sample (NAKO).

| Study | Group | N | Mean Age | Sex |
| --- | --- | --- | --- | --- |
| ADNI | HC | 1073 | $68.36 \pm 3.3$ | 634 females (59.09%) |
| | MCI | 1529 | $66.71 \pm 4.25$ | 729 females (47.67%) |
| | AD | 588 | $67.2 \pm 4.65$ | 291 females (49.48%) |
| AIBL | HC | 368 | $68.00 \pm 2.77$ | 217 females (58.97%) |
| | MCI | 78 | $68.05 \pm 3.54$ | 33 females (42.31%) |
| | AD | 28 | $66.89 \pm 4.44$ | 16 females (57.14%) |
| OASIS3 | HC | 1643 | $63.36 \pm 6.85$ | 1028 females (62.57%) |
| | MCI | 63 | $66.67 \pm 4.85$ | 37 females (58.73%) |
| | AD | 228 | $66.54 \pm 4.94$ | 97 females (42.54%) |
| NIFD | HC | 263 | $62.71 \pm 6.41$ | 148 females (56.27%) |
| | FTD | 317 | $63.26 \pm 5.66$ | 120 females (37.85%) |
| MACS | HC | 835 | $35.71 \pm 12.6$ | 528 females (63.23%) |
| NAKO | HC | 29883 | $48.45 \pm 12.09$ | 13201 females (44.18%) |

## Appendix B Sensitivity Analysis

To establish the impact of the hyperparameter *k* in our application of the N^3^ framework, we have added a sensitivity analysis and repeated the statistical and machine learning analyses for *k* ∈ [5, 35].

The sensitivity analysis demonstrates robust N^3^ performance across the tested parameter range, with performance metrics exhibiting minimal variation for Δ*k <* 15. We see two general observable trends: First, values for *k >* 20 tend to decrease performance and second, with increasing *k*, the trade-off between recall and precision is shifted towards higher recall.

The sensitivity analysis revealed disease-specific optimal configurations in group-level analyses (see Table B2 and Fig. B1). Statistical discrimination peaks at k=5 for AD (*η*^2^=0.299), k=15 for MCI (*η*^2^=0.067) and FTD (*η*^2^=0.376), reflecting differential spatial scales of pathological alterations. For AD, statistical discrimination declines monotonically with increasing k, suggesting that fine-grained local density estimation optimally captures AD-related brain alterations. Conversely, FTD exhibits peak discrimination at k=15, indicating that moderately larger neighborhoods better characterize frontotemporal pathology patterns. MCI shows relatively stable but modest effect sizes across *k* ∈ [10, 20] (*η*^2^0.064).

Classification performance too exhibited disease-dependent optimal *k* values (see Table B3 and Fig. B2). AD achieves peak balanced accuracy at *k* ≈ 10 − 15 (0.791) with a high F1-score (0.631), representing a good trade-off between precision and recall. FTD classification shows a precision-recall trade-off, while *k* = 5 maximizes recall (0.805), *k* = 25 yields highest precision (0.880) and balanced accuracy (0.786). MCI demonstrates consistent but modest classification performance (balanced accuracy 0.610–0.620), with marginal advantage at k=15 (0.620).

The discordance between optimal *k* for statistical discrimination versus classification, particularly evident in FTD and MCI, reflects the distinction between group-level distribution shifts and individual-level decision boundary optimization.

**Fig. B1.**
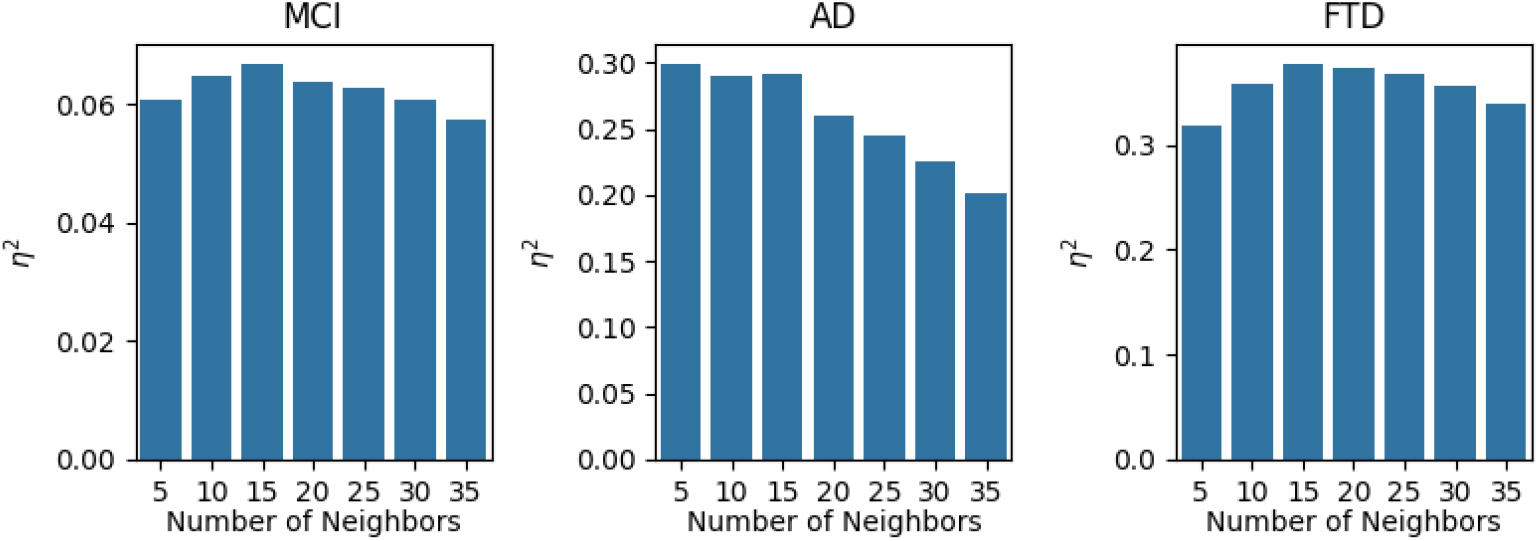
Statistical effect sizes for different values of the nearest neighbor parameter *k*. Effect size, partial eta-squared (*η*^2^), is depicted, comparing the discriminative power of the N^3^ biomarker across neurodegenerative diseases for different values of the nearest neighbor parameter *k* (MCI - Mild Cognitive Impairment, AD - Alzheimer’s Disease, FTD - Frontotemporal Dementia).

## Appendix C Normative Modeling Aggregation

We evaluated different aggregation strategies of the Normative Modeling z-scores. The tables summarize effect sizes from group-level statistics and predictive performance from machine learning analyses, allowing a direct comparison of how each aggregation strategy supports disease classification. Aggregations based on the overall count of outliers and the absolute sum of deviations yielded consistently larger effect sizes in the statistical analyses, whereas averaging and the UMAP-based aggregation of z-scores resulted in negligible effects for MCI and AD. In contrast, machine learning classifiers benefited from UMAP-based aggregation, particularly in FTD, and from averaging in MCI, indicating that these strategies encode informative structure that can be exploited by non-linear models for case-level prediction. Overall, different diseases benefit from different aggregation strategies, while the count of outliers emerges as a robust choice for both statistical and machine learning analyses across diseases.

**Table B2.** Statistical sensitivity analysis of nearest neighbor parameter *k*. Effect of the nearest neighbor parameter *k* on the discriminative power of the N^3^ biomarker across neurodegenerative diseases (MCI - Mild Cognitive Impairment, AD - Alzheimer’s Disease, FTD - Frontotemporal Dementia). F-statistics and partial eta-squared (*η*^2^) effect sizes comparing patient groups to cognitively unimpaired individuals. Bold indicates optimal results per disease. All p*<*0.001.

| $k$ | MCI | | AD | | FTD | |
| --- | --- | --- | --- | --- | --- | --- |
| | F | $\eta^2$ | F | $\eta^2$ | F | $\eta^2$ |
| 5 | F(1,4565)=296 | 0.061 | F(1,3709)=**1,577** | **0.299** | F(1,580)=269 | 0.318 |
| 10 | F(1,4565)=312 | 0.065 | F(1,3709)=1,510 | 0.290 | F(1,580)=321 | 0.358 |
| 15 | F(1,4565)=**313** | **0.067** | F(1,3709)=1,412 | 0.292 | F(1,580)=**346** | **0.376** |
| 20 | F(1,4565)=311 | 0.064 | F(1,3709)=1,302 | 0.260 | F(1,580)=342 | 0.373 |
| 25 | F(1,4565)=306 | 0.063 | F(1,3709)=1,198 | 0.244 | F(1,580)=335 | 0.367 |
| 30 | F(1,4565)=295 | 0.061 | F(1,3709)=1,074 | 0.225 | F(1,580)=319 | 0.356 |
| 35 | F(1,4565)=279 | 0.058 | F(1,3709)=932 | 0.201 | F(1,580)=296 | 0.339 |

**Fig. B2.**
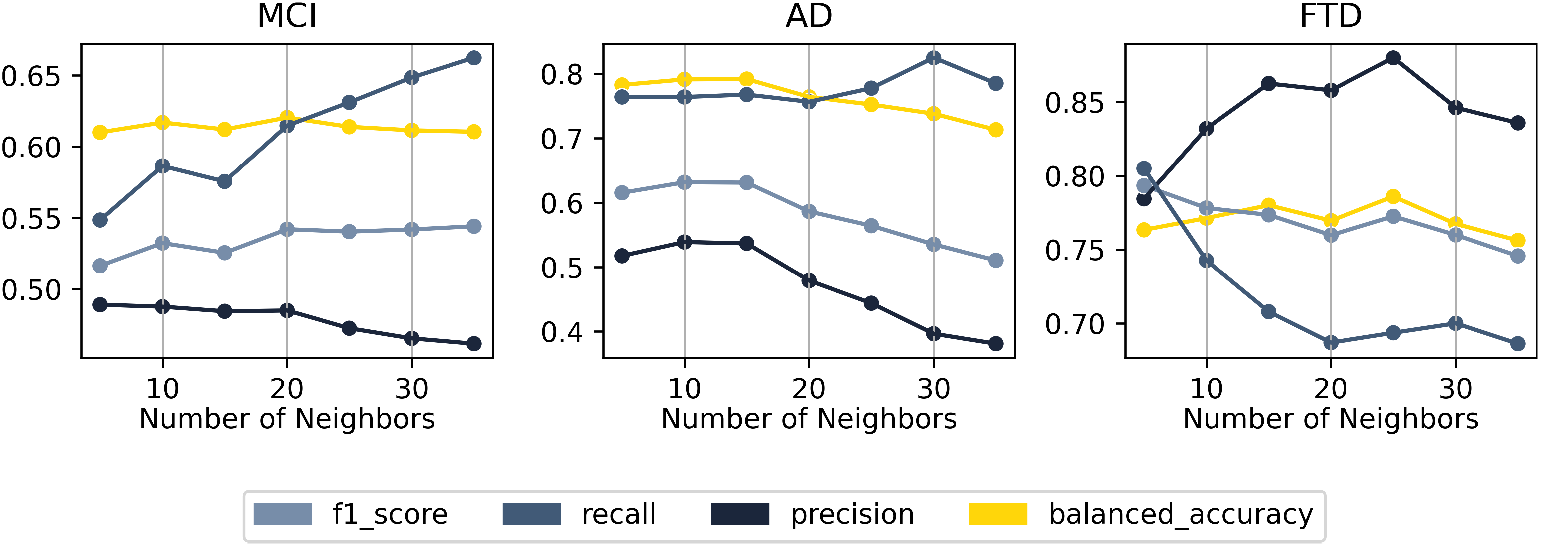
Machine Learning performance for different values of the nearest neighbor parameter. *k*. Performance metrics for the N^3^ biomarker for different values of the nearest neighbor parameter *k* in the classification of cognitively unimpaired individuals from those with neurodegenerative diseases (MCI - Mild Cognitive Impairment, AD - Alzheimer’s Disease, FTD - Frontotemporal Dementia).

**Table B3.** Classification performance for different values for the nearest neighbor parameter *k*. Bold values indicate best performance per disease and metric.MCI - Mild Cognitive Impairment, AD - Alzheimer’s Disease, FTD - Frontotemporal Dementia; Bal. Acc. - Balanced Accuracy. Bold indicates best performance per disease.

| Disease | k | Bal. Acc. | Recall | Precision | F1 Score |
| --- | --- | --- | --- | --- | --- |
| MCI | 5 | 0.610 | 0.548 | **0.489** | 0.516 |
| MCI | 10 | 0.617 | 0.586 | 0.488 | 0.532 |
| MCI | 15 | 0.612 | 0.576 | 0.484 | 0.526 |
| MCI | 20 | **0.620** | 0.615 | 0.485 | 0.542 |
| MCI | 25 | 0.614 | 0.631 | 0.473 | 0.540 |
| MCI | 30 | 0.611 | 0.648 | 0.465 | 0.542 |
| MCI | 35 | 0.610 | **0.662** | 0.462 | **0.544** |
| AD | 5 | 0.783 | 0.764 | 0.517 | 0.615 |
| AD | 10 | 0.791 | 0.764 | **0.539** | **0.632** |
| AD | 15 | **0.792** | 0.768 | 0.537 | 0.631 |
| AD | 20 | 0.764 | 0.757 | 0.479 | 0.586 |
| AD | 25 | 0.752 | 0.778 | 0.444 | 0.564 |
| AD | 30 | 0.738 | **0.825** | 0.396 | 0.535 |
| AD | 35 | 0.713 | 0.785 | 0.381 | 0.510 |
| FTD | 5 | 0.763 | **0.805** | 0.784 | **0.793** |
| FTD | 10 | 0.771 | 0.743 | 0.832 | 0.778 |
| FTD | 15 | 0.780 | 0.708 | 0.863 | 0.773 |
| FTD | 20 | 0.770 | 0.687 | 0.858 | 0.760 |
| FTD | 25 | **0.786** | 0.694 | **0.880** | 0.773 |
| FTD | 30 | 0.767 | 0.700 | 0.846 | 0.760 |
| FTD | 35 | 0.756 | 0.686 | 0.836 | 0.746 |

**Table C4.** Statistical discrimination of the different neurodegenerative diseases for different aggregation strategies of the normative modeling z-scores.

| Aggregation | MCI |  | AD |  | FTD |  |
| --- | --- | --- | --- | --- | --- | --- |
| | F | $\eta^2$ | F | $\eta^2$ | F | $\eta^2$ |
| average | F(1,4565)=1.45 | 0.0003 | F(1,3709)=0.16 | 0.0000 | F(1,580)=0.14 | 0.0002 |
| count outliers | F(1,4565)=74 | 0.0159 | F(1,3709)=1,073 | 0.223 | F(1,580)=119 | 0.173 |
| absolute sum | F(1,4565)=83 | 0.0180 | F(1,3709)=969 | 0.207 | F(1,580)=123 | 0.176 |
| UMAP | F(1,4565)=0.02 | 0.0000 | F(1,3709)=174 | 0.0449 | F(1,580)=149 | 0.206 |

**Table C5.**
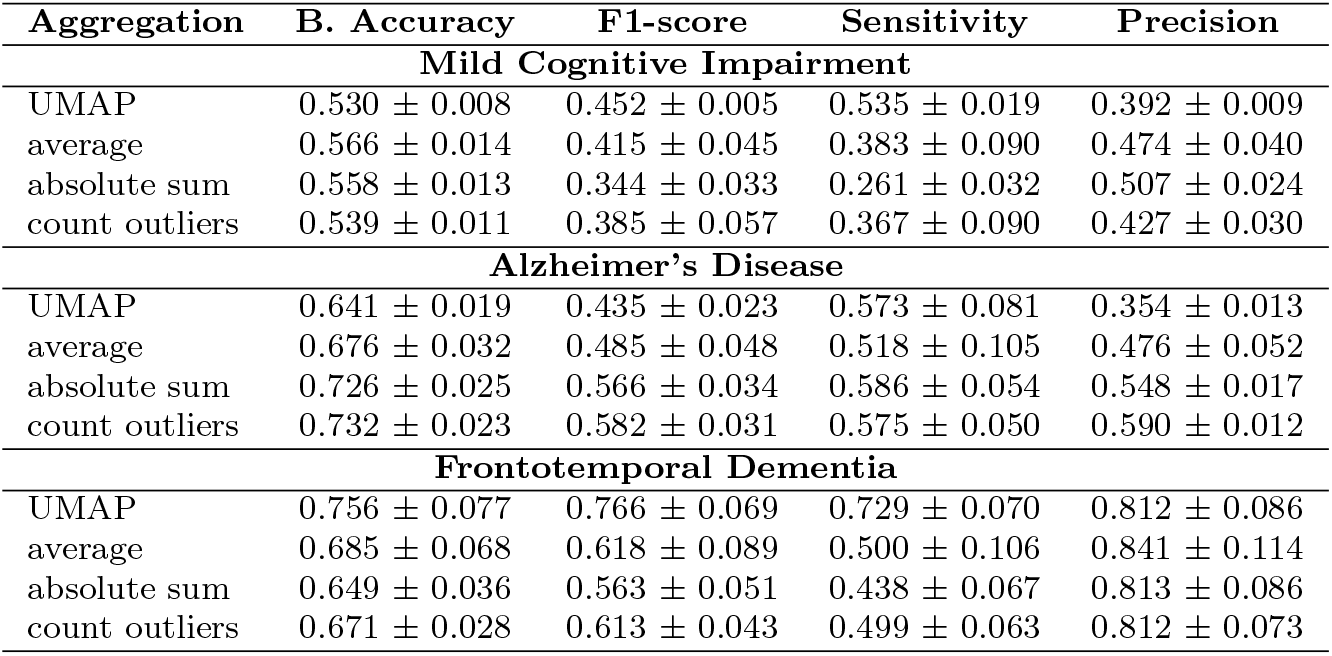
Performance metrics of the machine learning analyses for different aggregation strategies of the normative modeling z-scores.

| Aggregation | B. Accuracy | F1-score | Sensitivity | Precision |
| --- | --- | --- | --- | --- |
| **Mild Cognitive Impairment** |  |  |  |  |
| UMAP | $0.530 \pm 0.008$ | $0.452 \pm 0.005$ | $0.535 \pm 0.019$ | $0.392 \pm 0.009$ |
| average | $0.566 \pm 0.014$ | $0.415 \pm 0.045$ | $0.383 \pm 0.090$ | $0.474 \pm 0.040$ |
| absolute sum | $0.558 \pm 0.013$ | $0.344 \pm 0.033$ | $0.261 \pm 0.032$ | $0.507 \pm 0.024$ |
| count outliers | $0.539 \pm 0.011$ | $0.385 \pm 0.057$ | $0.367 \pm 0.090$ | $0.427 \pm 0.030$ |
| **Alzheimer's Disease** |  |  |  |  |
| UMAP | $0.641 \pm 0.019$ | $0.435 \pm 0.023$ | $0.573 \pm 0.081$ | $0.354 \pm 0.013$ |
| average | $0.676 \pm 0.032$ | $0.485 \pm 0.048$ | $0.518 \pm 0.105$ | $0.476 \pm 0.052$ |
| absolute sum | $0.726 \pm 0.025$ | $0.566 \pm 0.034$ | $0.586 \pm 0.054$ | $0.548 \pm 0.017$ |
| count outliers | $0.732 \pm 0.023$ | $0.582 \pm 0.031$ | $0.575 \pm 0.050$ | $0.590 \pm 0.012$ |
| **Frontotemporal Dementia** |  |  |  |  |
| UMAP | $0.756 \pm 0.077$ | $0.766 \pm 0.069$ | $0.729 \pm 0.070$ | $0.812 \pm 0.086$ |
| average | $0.685 \pm 0.068$ | $0.618 \pm 0.089$ | $0.500 \pm 0.106$ | $0.841 \pm 0.114$ |
| absolute sum | $0.649 \pm 0.036$ | $0.563 \pm 0.051$ | $0.438 \pm 0.067$ | $0.813 \pm 0.086$ |
| count outliers | $0.671 \pm 0.028$ | $0.613 \pm 0.043$ | $0.499 \pm 0.063$ | $0.812 \pm 0.073$ |

## Notes

### Competing Interest Statement

The authors have declared no competing interest.

### Funding Statement

This work was funded by the German Research Foundation (DFG grants HA7070/2-2, HA7070/3, and HA7070/4 to T.H.), the Interdisciplinary Center for Clinical Research (IZKF) of the medical faculty of Muenster (grants 22 Dan3/012/17 to U.D. and MzH 3/020/20 to T.H.) and the IMF research instrument of the medical faculty of Muenster (grant LE112409 to R.Leenings). X. Jiang was supported by the Deutsche Forschungsgemeinschaft (DFG) under Grant CRC 1450-431460824.
This project was conducted with data from the German National Cohort (NAKO) (www.nako.de). The NAKO is funded by the Federal Ministry of Education and Research (BMBF) [project funding reference numbers: 01ER1301A/B/C, 01ER1511D, 01ER1801A/B/C/D and 01ER2301A/B/C], federal states of Germany and the Helmholtz Association, the participating universities and the institutes of the Leibniz Association. We thank all participants who took part in the NAKO study and the staff of this research initiative.
Data collection and sharing for this project was funded by the Alzheimers Disease Neuroimaging Initiative (ADNI) (National Institutes of Health Grant U01 AG024904) and DOD ADNI (Department of Defense award number W81XWH-12-2-0012). ADNI is funded by the National Institute on Aging, the National Institute of Biomedical Imaging and Bioengineering, and through generous contributions from the following: AbbVie, Alzheimer's Association; Alzheimer's Drug Discovery Foundation; Araclon Biotech; BioClinica, Inc.; Biogen; Bristol-Myers Squibb Company; CereSpir, Inc.; Cogstate; Eisai Inc.; Elan Pharmaceuticals, Inc.; Eli Lilly and Company; EuroImmun; F. Hoffmann-La Roche Ltd and its affiliated company Genentech, Inc.; Fujirebio; GE Healthcare; IXICO Ltd.; Janssen Alzheimer Immunotherapy Research Development; LLC.; Johnson Johnson Pharmaceutical Research Development LLC.; Lumosity; Lundbeck; Merck Co., Inc.; Meso Scale Diagnostics, LLC.; NeuroRx Research; Neurotrack Technologies; Novartis Pharmaceuticals Corporation; Pfizer Inc.; Piramal Imaging; Servier; Takeda Pharmaceutical Company; and Transition Therapeutics.
The Canadian Institutes of Health Research is providing funds to support ADNI clinical sites in Canada. Private sector contributions are facilitated by the Foundation for the National Institutes of Health (www.fnih.org). The grantee organization is the Northern California Institute for Research and Education, and the study is coordinated by the Alzheimer's Therapeutic Research Institute at the University of Southern California. ADNI data are disseminated by the Laboratory for Neuro Imaging at the University of Southern California.
Data was in part provided by OASIS-3: Principal Investigators: T. Benzinger, D. Marcus, J. Morris; NIH P50AG00561, P30NS09857781, P01AG026276, P01AG003991, R01AG043434, UL1TR000448, R01EB009352. AV-45 doses were provided by Avid Radiopharmaceuticals, a wholly owned subsidiary of Eli Lilly.
Data collection and sharing for this project was funded by the Frontotemporal Lobar Degeneration Neuroimaging Initiative (National Institutes of Health Grant R01 AG032306). The study is coordinated through the University of California, San Francisco, Memory and Aging Center. FTLDNI data are disseminated by the Laboratory for Neuro Imaging at the University of Southern California. The Investigators at NIFD/FTLDNI contributed to the design and implementation of FTLDNI and/or provided data, but did not participate in analysis or writing of this report (unless otherwise listed). The FTLDNI investigators included the following individuals: Howard Rosen; University of California, San Francisco (PI) Bradford C. Dickerson; Harvard Medical School and Massachusetts General Hospital Kimoko Domoto-Reilly; University of Washington School of Medicine David Knopman; Mayo Clinic, Rochester, Bradley F. Boeve; Mayo Clinic Rochester Adam L. Boxer; University of California, San Francisco, John Kornak; University of California, San Francisco Bruce L. Miller; University of California, San Francisco William W. Seeley; University of California, San Francisco Maria-Luisa Gorno-Tempini; University of California, San Francisco Scott McGinnis; University of California, San Francisco Maria Luisa Mandelli; University of California, San Francisco

### Author Declarations

The study used data from the MACS cohort, which was approved by the ethics committees of the medical faculties of the University of Marburg (07/2014) and the University of Muenster (2014-422-b-S). All procedures were performed in accordance with ethical guidelines and regulations. Participants provided written informed consent before examination and received financial compensation for participation. This study furthermore uses publicly available study data. Please see the study-specific documentations. (NAKO: nako.de/forschung, ADNI, AIBL, NIFD: ida.loni.usc.edu, OASIS3: sites.wustl.edu/oasisbrains)

### Summary of Updates

Streamlined Introduction, Results and Discussion. Introduced further conceptual background. Revised Algorithm Description. Additional Sensitivity Analyses.

